# Efficacy of a parent-child program for 3- to 6-year-old children with stuttering: a retrospective controlled wait-list group pilot study

**DOI:** 10.1101/2025.06.20.25329968

**Authors:** Nicole E. Neef, Imke Niemann, Anna Merkel, Kristina Anders, Katja Hente, Johanna Margarete Joisten, Alexander Wolff von Gudenberg, Christian H. Riedel

**Author notes:** Corresponding author: Nicole Neef.

## Abstract

We evaluated the efficacy of Frankini, a 12-month early parent-child intervention that combines online parent counseling with hybrid speech restructuring to reduce stuttering severity and promote fluency-supportive interaction.

This retrospective, nonrandomized pilot trial included cases enrolled between September 2019 and November 2023. For analysis, only participants who completed Module 1 (indirect parental training) and Module 2 (the first hybrid speech restructuring module) were included. A total of 51 cases met all inclusion criteria, and 30 of these completed all three modules.

To simulate a wait-list-controlled design, eligible participants were divided into early and delayed groups using median split. The early group completed Module 2 nine months after baseline, the delayed group twelve months after baseline. Groups were matched on key characteristics and differed only in the timing of the first direct intervention. Blinded raters assessed stuttering severity.

Primary outcomes included the Stuttering Severity Index, parental severity rating, and a 10-item parent report. At 9 months, the early group showed reduced stuttering severity, while the delayed group showed no change (mean difference = -8.33 95%CI [-12.98, -3.68], p < 0.001, with d = -1.14). By 12 months, both groups improved, and group difference were no longer significant (mean difference = -3.37 95%CI [-8.23, 1.50], p = 0.168 and d = -0.48). Parental ratings mirrored these outcomes showing consistent improvement after each module.

Speech restructuring significantly improved speech fluency and parent counseling enhanced parents’ confidence, supporting the value of initiating treatment before age 6; however, follow-up is needed to assess long-term effects.

**Trial Registration:** <u>DRKS00034731</u>.

## Introduction

Approximately 5% of children experience stuttering (Bloodstein et al., 2021). Overt symptoms include sound and syllable repetitions, sound prolongations, and speech blocks ─ involuntary disruptions that the speaker perceives as a loss of control ─ often resulting in struggle, compensatory behaviors, and a negative impact on life (Tichenor & Yaruss, 2021). Stuttering typically begins in early childhood, with an onset ranging from 2 to 5 years and a mean age of 36 months (Yairi & Ambrose, 2013). Spontaneous remission occurs in 75% of cases within the first 18 months (Reilly et al., 2009; Yairi & Ambrose, 2013), however no marker currently predicts whether stuttering will persist. Despite a high remission rate, 1% of the population continues to stutter throughout life (Yairi & Ambrose, 2013). A recent Cochrane review identified early treatment as the most effective treatment (Sjøstrand et al., 2021), and because of the high neuroplasticity in young children (Neef & Chang, 2024), early interventions are highly desirable. Treatment approaches for preschoolers who stutter are recommended in Germany (Katrin Neumann et al., 2017), however, referrals for therapy peak late, at 6 years of age (Sommer et al., 2021), and efficacy studies are rare (Brignell et al., 2021; Sjøstrand et al., 2021), with only one conducted in German preschoolers (Lattermann et al., 2008).

Worldwide, there are four treatment approaches for preschoolers who stutter that have been shown to reduce stuttering and improve communication attitudes in young children: The Lidcombe Program (Harris et al., 2002; Jones et al., 2005; Lattermann et al., 2008; Lewis et al., 2008; Onslow et al., 2003), the Palin Parent-Child Interaction therapy (Millard et al., 2009, 2018; Preston et al., 2022), the RESTART Demands and Capacities Model (Sonneville-Koedoot et al., 2015), and the combined fluency rules program (FRP) with parent-child interaction training (Bafrooei et al., 2025). The four approaches are broadly comparable in effectiveness but differ in methodology and focus, offering flexibility for tailoring treatment to the needs of the child and family. The Lidcombe Program is a direct therapy that explicitly focuses on reducing stuttering by reinforcing fluent speech and correcting stuttered speech through verbal contingencies. Parents deliver therapy in everyday speaking situations guided by a trained clinician (Onslow et al., 1994). Palin PCI and RESTART-DCM are mainly indirect therapies. Palin-PCI (Kelman & Nicholas, 2019) focuses on enhancing parent-child interaction and reducing environmental demands to support fluency, using video analysis, tailored feedback, and collaborative goal-setting with parents. If the child continues to show concern about their stuttering, therapy may be extended to include more direct strategies, such as increasing the child’s understanding of speech and communication and helping them develop personalized cognitive strategies for managing stuttering (Franken et al., 2022). RESTART-DCM (Franken et al., 2025) aims to balance speaking demands with a child’s capacities by reducing environmental pressure and supporting parental involvement, enabling the child to communicate with greater ease, with or without stuttering. In its first phase, the program focuses on reducing motor, linguistic, emotional, and cognitive demands arising within the child and from the environment. In the subsequent phase, the child works more directly on strengthening these underlying capacities; however, this phase does not directly target stuttering or fluency (Franken et al., 2025). The combined FRP with parent-child interaction training is a flexible stuttering intervention that promotes speech control through self-monitoring, individualized feedback, and non-verbal cues targeting both disfluencies and associated behaviors together with parent education and coaching to support fluency-enhancing strategies (Bafrooei et al., 2025). All four approaches require active parental involvement, but the intensity and nature of the role vary. Lidcombe involves structured verbal feedback from parents; and Palin PCI and RESTART-DCM focus on modifying interaction styles and environments rather than targeting stuttering directly. The most recent trial combined both modifying stuttering and educating parents about stuttering and parent-child interactions.

Thus, therapy for preschool children who stutter generally follows one of two complementary approaches: parent-focused or child-focused. Parent-focused (indirect) therapy aims to equip caregivers with strategies to foster a supportive, low-pressure communication environment. Child-focused (direct) therapy, on the other hand, involves teaching children techniques to manage and modify their stuttering directly. Each approach offers unique benefits and presents certain considerations. While both depend on active parental involvement, tailoring the level of engagement to individual family circumstances can enhance feasibility and success. Direct approaches introduce children to practical fluency-enhancing tools, such as easy onsets and slowed speech, and can empower them to develop awareness and resilience around their speech patterns. However, it’s important to match the intervention to a child’s developmental readiness, as younger children may need more support in integrating these strategies. For some, focusing explicitly on speech may increase self-monitoring; with careful guidance, this can be managed in a way that builds confidence rather than anxiety. Crucially, the field continues to refine its understanding of what works best for whom. While promising outcomes have been observed with both models (Brignell et al., 2021; K. Neumann et al., 2016; Katrin Neumann et al., 2017; Sjøstrand et al., 2021), further research is needed to explore the long-term impact of these strategies, individually and in combination. The diversity among children, differences in temperament, stuttering severity, family dynamics, and cultural background, underscores the value of flexible, individualized therapy planning (Millard et al., 2018). Finally, the evolving definitions of therapeutic success, ranging from fluency gains to increased communicative confidence and quality of life, reflect the broader, more holistic view emerging in the field (Brundage et al., 2021). This diversity of goals enriches clinical practice and encourages ongoing dialogue about best practices in early stuttering intervention.

Here, we assessed the efficacy of Frankini, an early intervention program designed for parents and preschoolers, that combines online parent counseling and training with speech restructuring techniques delivered both in-person and via telehealth. The program aimed to reduce stuttering severity while fostering interaction styles that promote fluent communication. By integrating both indirect and direct therapeutic elements, Frankini seeks to address the multifaceted nature of early childhood stuttering; that is supporting the child’s speech development while empowering caregivers to create a communicatively supportive home environment. The hybrid delivery format enhances accessibility for families who may face logistical or geographical barriers to in-person services, and allows for continuity of care across settings. This approach reflects a growing recognition of the importance of early, comprehensive intervention that aligns with both the child’s speech capacities and the family’s resources and dynamics. Importantly, Frankini does not equate therapeutic success solely with fluency gains; rather, it combines fluency-shaping strategies with an emphasis on acceptance, resilience, and communicative confidence, thereby resonating with the broader, neurodiversity-informed perspective increasingly valued in the stuttering field. The program is an adaptation of the Kasseler Stuttering Therapy, a computer-assisted fluency-shaping program developed for adults who stutter (Euler et al., 2009; Wolff-von-Gudenberg & Euler, 2000). Previously, we introduced an adoption of this program for school-aged children (FRANKA, (Euler et al., 2021). The new program, Frankini, combines indirect and direct treatment components across three modules spread over the course of 12 months (Figure 1A). Since spontaneous remission is still possible between the ages of three and six, and the course of stuttering often fluctuates, parents and therapists decide together after each module whether to continue, pause, or end treatment. After Module 1, key indicators for initiating intensive in-person therapy included the level of distress experienced by the child and parents, the severity of stuttering, and the prognosis for its progression.

**Figure 1.**
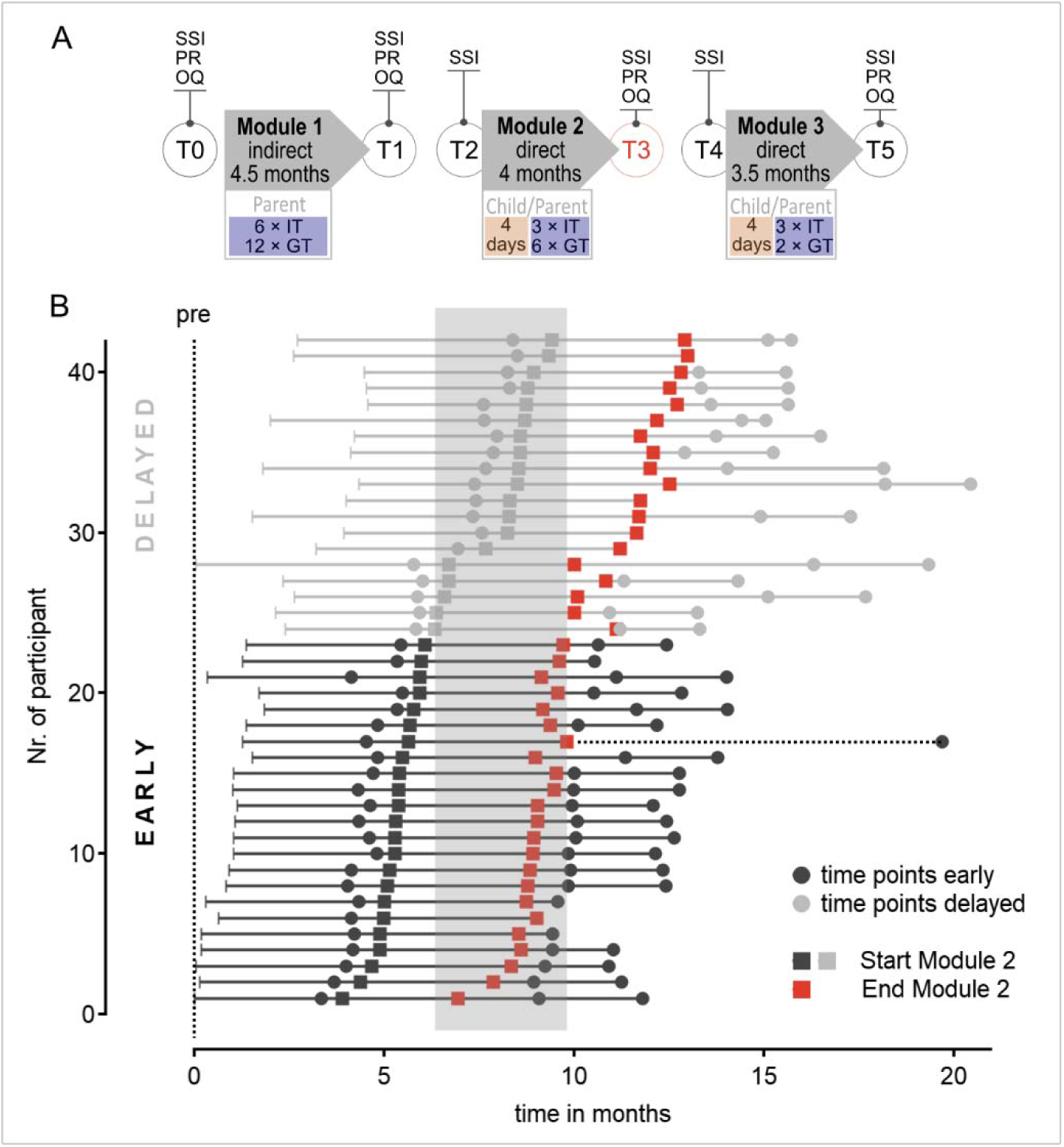
Procedure of the parent child program and participant’s group allocation. A) T0 – T5 are assessment time points. Assessments included Stuttering Severity Index (SSI), subjective parent rating (PR), online questionnaire on parents’ experience with the child’s stuttering (OQ). Modules included online sessions (blue) therapy in presence (red), individual therapy (IT) and group therapy (GT). B) Individual time schedules and group assignments of the participants.

Randomized controlled trials of behavioral interventions in neurodevelopmental disorders are notoriously difficult to conduct. Recruiting and retaining families is difficult because of the time-intensive nature of participation, especially for those already burdened by the disorder. The required long-term commitment often leads to high dropout rates, affecting study validity. Withholding treatment from the control groups raises ethical concerns, particularly when early intervention is crucial. Additionally, blinding is challenging because parents and clinicians often know if an intervention is active. We present an alternative approach by simulating a waitlist group design through the retrospective allocation of participants into early and delayed groups. Waiting times after registration varied, as parents were assigned to courses based on availability and compatibility with the family’s schedule. At nine months post-baseline, the early group had completed a direct treatment module, while the delayed group had not yet started the same module, and an interaction between group and time would indicate an effect of the intervention. Six recordings of speech samples throughout the program allowed for the comparison of objectively quantified stuttering severity at this relative point in time. This study design accounted for maturation processes and spontaneous remission, ensuring that the likelihood of these influences was equally distributed across both groups. Additionally, raters of the stuttering severity assessment were blinded to the time point of recording, reducing bias related to expectations of progress over time.

To our knowledge, this is the first study to apply a retrospective simulated wait-list design. In contrast to a prospective wait-list control group, where participants are knowingly delayed in receiving treatment, this design uses pre-existing clinical data to simulate a control group, with all outcome measures collected before group assignment. This approach offers distinct ethical and practical advantages, particularly in early childhood stuttering intervention, where timely access to treatment is critical and delays may contribute to emotional strain and frustration for both children and their families. By avoiding intentional withholding of therapy, the design reduces the risk of demotivation, dropout, and dissatisfaction that can occur in families placed on a traditional wait list. At the same time, the retrospective nature of the design entails certain methodological limitations, such as the absence of randomization and the potential for selection bias. Nonetheless, it provides a feasible and ethically responsible alternative for evaluating intervention effects in real-world clinical settings where randomized controlled trials may be difficult to implement.

Despite promising evidence from existing approaches, there is currently no evidence on the efficacy of the Frankini method. To address this gap, the present study applied a retrospective simulated wait-list design to evaluated the efficacy of this novel hybrid intervention program for preschool children who stutter. Frankini integrates telehealth coaching of parents with both individual and group sessions delivered online and in person to support children and their families. We examined whether the program leads to reductions in stuttering severity compared to no intervention, and whether parental confidence and coping improved over the course of treatment. We hypothesized that (1) children receiving direct intervention would show greater reductions in stuttering severity than those receiving no direct intervention, and (2) parents would report greater improvement in confidence and coping over time.

## Methods

The Frankini program combines telehealth coaching of parents with individual and group sessions delivered both online and in person, and its efficacy was evaluated using objectively measured stuttering severity, parent-reported severity ratings, and coping measures assessed through online surveys. This retrospective study was approved by the Ethical Review Board of the University Medical Center Göttingen and registered at the UMG study center (ID 2024-03288). The study protocol was registered in the German Clinical Trials Register https://www.drks.de/DRKS00034731.

### Participants Flow

The trial was a retrospective, two-armed, controlled, monocentric trial with a prospective simulated wait-list design. In *the retrospective phase,* eligible cases were identified from all 79 families who entered the program between September 2019 and November 2023. Families contacted us through various channels, including referrals from pediatricians and speech-language therapists, or through independent online searches. No formal exclusion criteria were applied at intake, other than age (3–6 years) and clinical indicators consistent with developmental stuttering. Eligibility required parental consent for repeated clinical and research assessments, video calls, daily video recordings, and storage and processing of pseudonymized data for treatment evaluation. No systematic sampling strategy was applied. Inclusion criteria required participation in Module 1 (indirect parent training) and Module 2 (first speech restructuring module). Families who paused after Module 1 or began directly with Module 2 were excluded. Additionally, four families discontinued without mutual agreement, one family transitioned to another therapy, and one child was withdrawn due to illness. As shown in the participant flow chart (Figure 2), 51 cases met the eligibility criteria.

**Figure 2.**
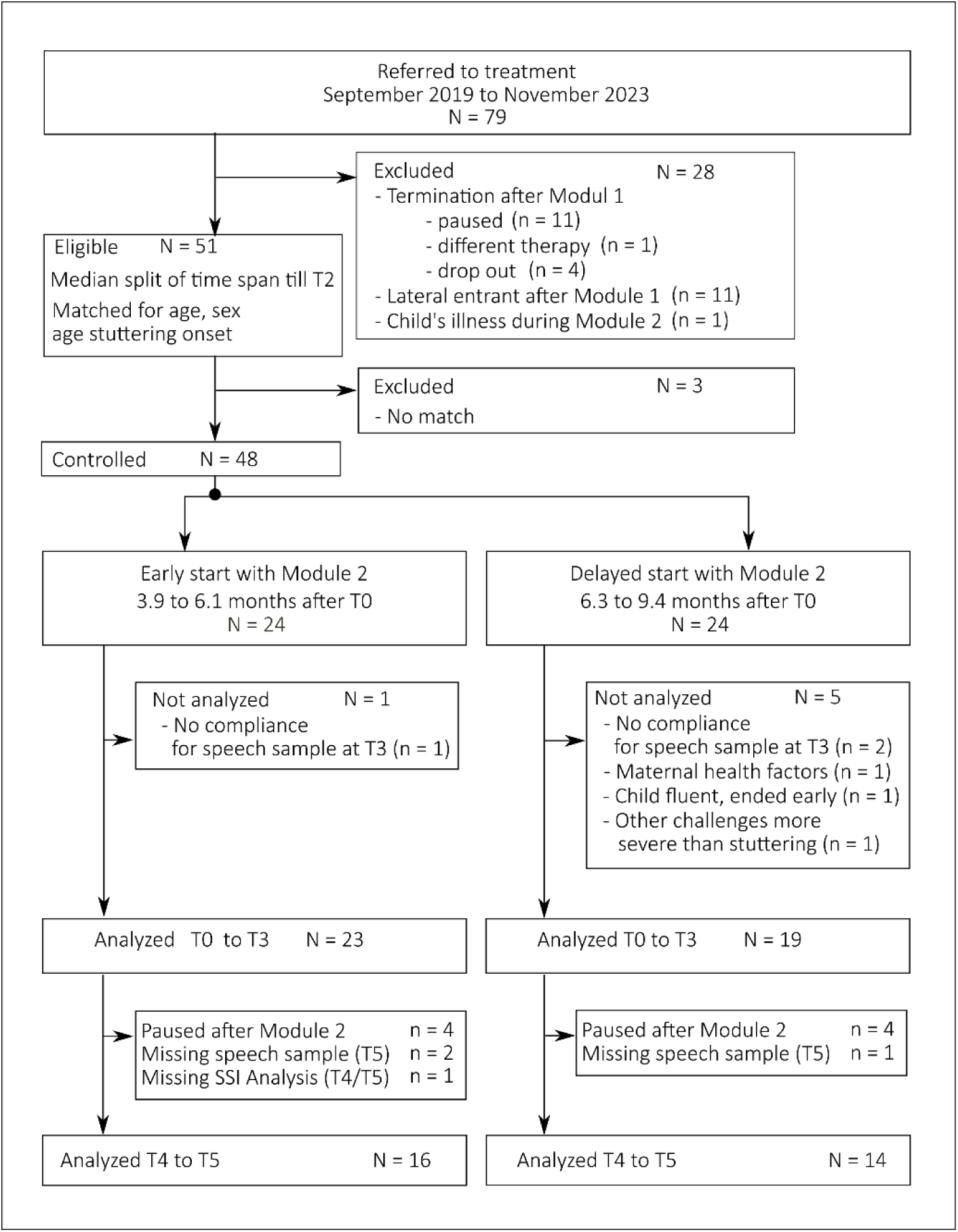
**Participant flow diagram** illustrates the sequence of identifying participants for inclusion in the analysis.

In *the prospective phase*, the 51 eligible cases identified during the retrospective phase were assigned to early or delayed intervention groups based on naturally occurring variation in the time between their initial clinical assessment (T0) and the completion of Module 2 (T2). This variation in treatment initiation allowed for a simulation of a waitlist-controlled design. Participants were divided into two groups using a median split of the time interval between T0 and T2 (Mdn = 6.3 months). The cutoff of 6.3 months was chosen because it represented the median waiting time among the 42 participating families. This approach ensured an even split between the shorter and longer waiting time groups and allowed for a balanced comparison The early group completed Module 2 on average within nine months, while the delayed group reached the same point after approximately twelve months (see Table 1, Figure 1B).

**Table 1.**
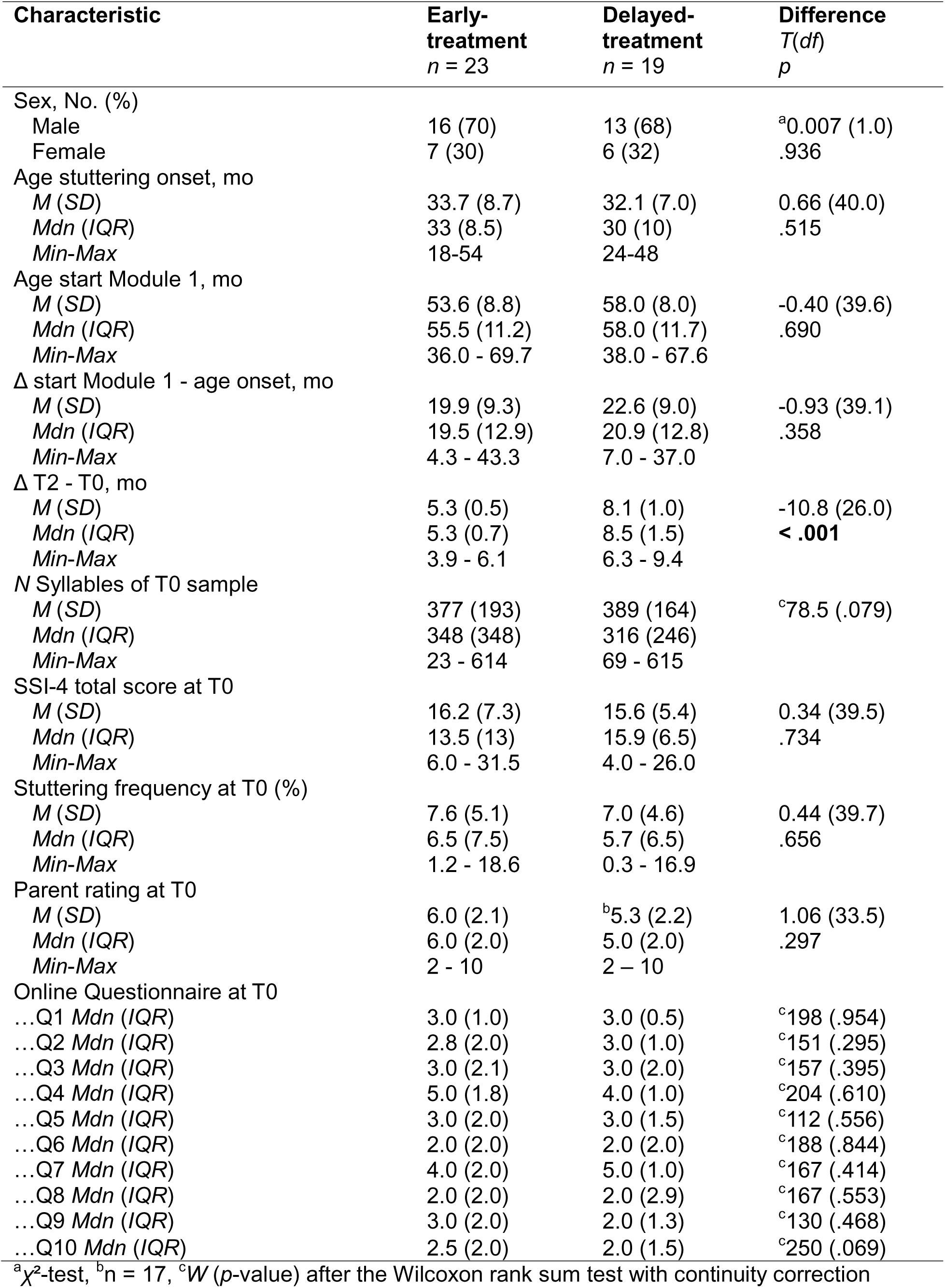
Baseline demographic and clinical characteristics.

To reduce potential confounds, group matching was performed on key variables: sex, age, and age of stuttering onset. During this process, three participants had to be excluded due to matching limitations, resulting in a final sample of 48 children, 24 in each group, who were included in the study analyses. SSI-4 ratings were then conducted by three blinded raters for the final matched sample. During this phase, some speech samples were found to be ineligible for analysis due to non-compliance. At the same time, reasons for missing samples were investigated for the first time, a documentation oversight on our part that could not be corrected retrospectively. As a result, valid SSI-4 data were obtained for 42 children (see Participant Flow Chart for details).

### Participants

Of the 42 children included in the analysis, 23 were assigned to the early group and 19 to the delayed group. Participants ranged in age from 3 to 5 years and included 29 boys and 13 girls. The mother tongue of 39 children was German. Among the 10 bilingual children, three had a different primary language, Albanian, English, or Russian, with German as their second L1 language. The remaining seven were native German speakers with a second L1 language, including English, Polish, Persian, or Russian.

A family history of stuttering was reported for 26 children, while 14 had no known family history; for 2 children, this information was unavailable. Twenty children had not received prior treatment for stuttering, while 22 had previously undergone some form of therapy. Information on family history, bilingualism, and prior treatment is summarized in Supplementary Table 1. Individual demographic data are presented in Supplementary Table 8.

At baseline (T0), stuttering severity as assessed by the SSI-4 was categorized as very mild in 10 children, mild in 15, moderate in 16, and severe in one case.

### Intervention

The Frankini program is a structured, year-long parent-child intervention for preschool-aged children who stutter (ages 3 - 6). It comprises three consecutive modules combining indirect and direct therapeutic elements, with parents actively involved throughout (Figure 1A). A description of the program has been published in a German clinical journal (Anders et al., 2023). Below we provide an overview of the intervention.

All therapists participating in this study were experienced members of the Kasseler Stottertherapie team (Euler et al., 2009, 2014, 2021; Wolff-von-Gudenberg & Euler, 2000) a specialized center for intensive stuttering treatment. Group sessions are co-led by two therapists to support consistency and collaborative delivery. As this was a pilot study, formal fidelity monitoring procedures (e.g., checklists, session recordings, or inter-rater reliability) were not yet implemented, and establishing fidelity was not a primary focus. However, informal fidelity was supported through internal supervision, shared structured materials, and the use of a common online therapy platform. These elements helped promote coherence across sessions and therapists. A formal treatment manual is currently in development to guide future studies, including a planned randomized controlled trial with more rigorous fidelity monitoring.

The program aims to promote speech confidence, reduce stuttering severity, and foster acceptance and openness around stuttering. Rather than enforcing fluency, the focus is on building communicative agency and early use of supportive strategies.

#### Module 1 - Parent-Only Online Training

Module 1 was delivered entirely online and focused on parent counseling and training, without direct child involvement. Through individual and small-group video sessions, parents received education on stuttering, developed a more confident and accepting stance, and learned to communicate openly and age-appropriately about stuttering. Practical strategies included structuring speaking situations, adjusting linguistic demands, and practicing soft voice onsets, an early fluency-shaping technique, through home-based exercises. These were practiced by parents alone before being introduced to children in later modules. Parents also submitted video recordings of interactions with their child, which were reviewed with therapists.

An integral part of this phase was the use of flunatic.mini software (Euler et al., 2009), which provides visual feedback on vocal onset to support the learning of soft voice onsets. Flunatic.mini is part of an online platform, that also offers written guidance, video demonstrations, fluency tracking tools, and reflection prompts.

#### Modules 2 and 3 - In-Person Parent-Child Intervention

Modules 2 and 3 each began with a four-day in-person phase involving group and individual parent-child sessions. In Module 2, many children encountered peers who stutter for the first time, helping to reduce stigma and encourage open discussion about stuttering. Children learned to identify and name stuttering symptoms using visual symbols in a supportive, pressure-free environment. Parents were present throughout and could step in during exercises if needed.

The core fluency technique, soft voice onset (“feather words”), was introduced through structured, play-based exercises that increased in linguistic complexity. Children practiced the technique in both group and individual settings, with reinforcement through flunatic.mini. Post-intensive phases included biweekly online sessions for individualized feedback.

Therapy was not aimed at achieving consistent use of soft onsets or prompting children to use them continuously. Instead, parents were guided to select appropriate home activities and adjust linguistic demands to match their child’s capabilities. Most children began practicing at the word or sentence level in Module 2, with Module 3 focusing on longer utterances and simple transfer tasks (e.g., ordering ice cream or sending a voice message). Maintaining speaking enjoyment remained central. Group participation also fostered learning by modeling, both from therapists and from peers, and helped desensitize children and parents to stuttering and fluency techniques.

#### Goals and Decision Points

Therapy goals across modules were tailored to each child’s developmental level and family context. These included reducing stuttering severity, enhancing communication confidence, increasing self-efficacy in both child and parent, and promoting a non-stigmatizing, open approach to stuttering. At the end of each module, therapists and parents jointly decided whether to continue, pause, or conclude therapy, based on observed progress, current symptom severity, family distress, and the potential for spontaneous recovery.

(Euler et al., 2009)The program courses between September 2019 and November 2023 involved five to eight families per course, with a total of 13 completed courses. The 13 courses were conducted by eight therapists, including the coauthors I.N., K.A., A.M., and K.H.

### Outcomes

The objective assessment of stuttering severity was conducted using the Stuttering Severity Index (SSI-4, (Riley, 2009), which was conducted at six time points: at the initial assessment before Module 1, at the end of Module 1, and at the beginning and end of the two hybrid modules (T0 - T5, Figure 1A). At each time point, a speech sample was recorded during a video call in which the therapist engaged the child in a conversation. Additionally, at T0, T3, and T5, parents provided videos of the children in everyday situations. Because speech samples were not available for every child at each time point, all available samples for a child were analyzed at each time point. The number of analyzed syllables varied among the children, with a median of 348 syllables in the early group and 316 syllables in the delayed group at T0 (Table 1). Parent ratings of stuttering severity (PR) and the online questionnaire on parents’ experiences with the child’s stuttering (OQ) were conducted before Module 1 and at the end of each module (T0, T1, T3, and T5; Figure 1A). Parent-reported measures were scored on a Likert scale.

The surveys assessed parents’ subjective perception of their child’s stuttering severity as well as their attitudes and feelings about stuttering. The severity rating involved evaluating the child’s current symptoms on a scale of one (no stuttering) to 10 (very severe). Additionally, five questions gauged parents’ confidence in handling and discussing stuttering (e.g., “I can handle my child’s stuttering calmly.”), while another five questions measured their emotional responses and concerns (e.g., “My child’s stuttering worries me.”). Responses were recorded on a scale ranging from one (do not agree at all) to five (completely agree).

### Blinded assessment of stuttering severity

K.A. pseudonymized the speech samples and assigned them to three raters, including the coauthor A.M., all of whom are clinical speech scientists. Initially, T0 samples were assessed to test for baseline differences between the groups, with none found (Table 1). Subsequently, the remaining samples (T1–T5) were randomized according to the time point for analysis. Each rater analyzed all samples from a given child. No rater was involved in the intervention for that child.

The objective assessment of stuttering severity was conducted using the SSI-4 (Riley, 2009), which involves analyzing the frequency of stuttered syllables, measuring the duration of the three longest stuttering events, and evaluating physical concomitants such as grimacing and head and limb movements. SSI-4 provides a total score that encompasses three subscales: frequency, duration, and physical concomitants.

### Statistical methods

To test the efficacy of the parent-child program, we fitted the SSI-4 total scores with a linear mixed-effects model using maximum likelihood estimation (Supplementary Table 2). The baseline model included sex and age as fixed-effects variables, and subject and time as random-effects variables. We tested whether the model fit improved by including (i) group (early versus delayed treatment group), (ii) time (pre-, 9 months post-T0, 12 months post-T0), and (iii) group-by-time interaction. Additionally, we set the following orthogonal and non-orthogonal contrasts: The contrast for the group tested whether the early treatment group had a greater reduction than the delayed group; contrasts for time tested (i) whether the 9-month condition showed smaller SSI values than the pre-condition, and (ii) whether the 12-month condition showed smaller SSI values than the pre-condition. Post hoc unpaired *t*-tests were conducted to assess group differences at 9 and 12 months after baseline. To assess the effect size, we calculated Cohen’s *d*. Additionally, we conducted one-sample *t*-tests to determine whether the reduction in stuttering, as reflected by the subjective parent rating score, was significant. These tests were performed after treatment Module 2 and Module 3.

To test whether the parents’ experience with the child’s stuttering improved, we calculated the Wilcoxon signed rank test with continuity correction between baseline scores and scores at T1, T3, and T5. Additionally, we plotted the difference in scores between these time points and the baseline to visualize individual changes.

To explore the association between reductions in stuttering severity and changes in the parents’ experiences of their child’s stuttering, we calculated Spearman correlations between the change in SSI total score (T3-T0) and the change in questionnaire scores (T3-T0).

### Interrater agreement

We tested interrater agreement among all three raters. Therefore, each rater analyzed one randomly chosen speech sample from each child. This resulted in a total of 42 speech samples per rater (19%). Krippendorff’s alpha (Hughes, 2021), along with the macro developed by Hayes for ordinal and interval data (Hayes & Krippendorff, 2007), was used to calculate interrater agreement for both frequency (interval scale) and SSI total score (ordinal scale). Additionally, we calculated the intraclass correlation (ICC) with a two-way model, type agreement, to provide the 95% confidence interval for the ICC population values. Statistical analysis was performed using R version 4.4.0 using *irr*, *multcomp*, *nlme*, *pastecs*, *stats,* and*, tidyr*.

## Results

### Objective measure

Our analysis showed that treatment led to a significant reduction in stuttering severity, as reflected in the effect of time, χ*²*(9) = 22.0, *p* < 0.001. This reduction differed between the early and delayed treatment groups, as reflected in a significant group effect, χ*²*(7) = 5.1, *p* < 0.024 (Figure 3A). Most importantly, the treatment × group interaction was also significant, χ*²*(11) = 10.6, *p* = 0.005, indicating that the extent of improvement in stuttering severity depended on the timing of treatment.

**Figure 3.**
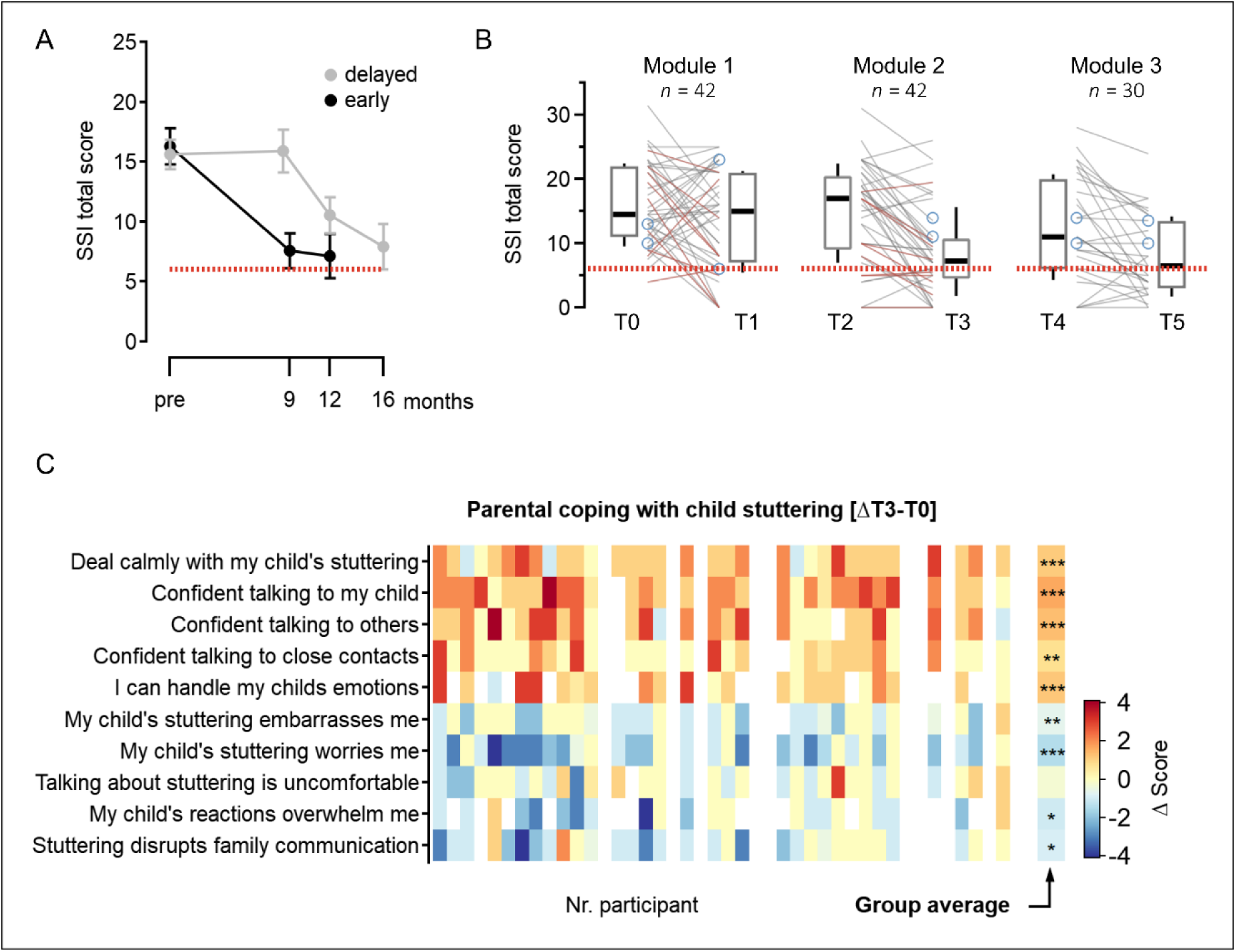
Changes in stuttering severity and parents’ experiences of their child’s stuttering over time. A) Stuttering severity (SSI) reduced after 9 months in the early treatment group, with no change in the delayed group. Plotted are group averages and standard errors. The dotted line represents the SSI cut-off value for stuttering. B) Box plots and individual curves for SSI total scores, separately plotted for the three treatment modules. Red lines indicate cases not participating in Module 3. Blue circles indicate cases with missing values. C) Heat map showing changes in parental experience with their child’s stuttering between baseline and T3 (after Module 2). White cells indicate missing data. The last column indicates the grand mean for a given item with \**p* < .05, \*\**p* < .01, and \*\*\**p* < .001 (Wilcoxon signed rank test with continuity correction).

By 9 months, children in the early treatment group showed a marked reduction in stuttering with a mean SSI-4 score of 7.6 (95% CI [4.6, 10.5]), a mean change of -8.7 points (95% CI [-12.2, -5.2]) from baseline. In contrast, the delayed group showed virtually no change during the same period, with a mean SSI-4 score of 15.9 (95% CI [12.1, 19.6]) and a negligible mean change of 0.3 (95% CI [-3.1, 3.6]). A post hoc unpaired *t*-test confirmed that the difference in SSI-4 total scores between the groups at 9 months post-baseline was statistically significant, with a mean difference of -8.33 (95% CI [-12.98, -3.68]), *t*(36.3) = -3.63, *p* < 0.001. This corresponds to a large effect size of *d* = -1.14 (95% CI [-1.82, -0.47]).

At the 12 month mark, both groups showed a reduction in stuttering severity. The early group manifested their improvement, with a mean score of 7.2 (95% CI [3.2, 11.1]), while the delayed group also improved to a mean score of 10.5 (95% CI [7.4, 13.7]). However, the difference between the groups at this point in time was no longer statistically significant (*t*(30.1) = -1.44, *p* = 0.168), although the effect size still suggested a moderate advantage for the early group (*d* = 0.49, 95% CI [-1.18, 0.21]). The mean change from baseline at 12 months was -10.6 (95% CI [-18.2, -3.3]) for the early group and -5.1 (95% CI [-8.0, -2.2]) for the delayed group. Supplementary Table 2 lists the details of the statistical analysis of the SSI total scores.

Additional analyses of percent stuttered syllables and SSI-4-subscores for frequency and duration confirmed these findings (Supplementary Tables 3 to 5). Group-wise mean changes in SSI-4 total scores from baseline (T0) to the end of Module 2 (T3) and Module 3 (T5) are reported in Table 2. Figure 3B shows the trajectory of stuttering severity over the course of the program for all 42 participants, regardless of the group assignment. Importantly, the most substantial improvements were observed after completion of Module 2, with gains largely maintained following Module 3 in children who continued the program.

**Table 2.** Summary of mean changes from baseline per group.

|  |  | **Early-treatment** |  | **Delayed-treatment** |  |
| --- | --- | --- | --- | --- | --- |
|  |  | at 9 months<br>*n* = 23 | at 12 months<br>*n* = 16 | at 9 months<br>*n* = 19 | at 12 months<br>*n* = 19 |
| Stuttered syllables % | *M* [*CI*<sub>95%</sub>]<br>*Mdn* (*Min* to<br>*Max*) | -5.3 [-7.3 to -<br>3.2]<br>-4.0 (-16.7 to<br>1.4) | -6.0 [-9.0 to -<br>3.1]<br>-5.1 (-15.4 to<br>3.5) | -0.8 [-3.6 to<br>1.9]<br>-0.8 (-11.7 to<br>7.6) | -3.8 [-5.4 to -<br>2.1]<br>-2.9 (-10.2 to<br>0.6) |
| SSI-4 total | *M* [*CI*<sub>95%</sub>]<br>*Mdn* (*Min* to<br>*Max*) | -8.7 [-12.2 to<br>-5.2]<br>-7.5 (-23.0 to<br>8.5) | -10.6 [-18.2<br>to -3.3]<br>17.0 (-31.5 to<br>18) | 0.3 [-3.1 to<br>3.6]<br>0.5 (-12.0 to<br>13.0) | -5.1 [-8.0 to -<br>2.2]<br>-6.0 (-14.5 to<br>4) |
| Frequency | *M* [*CI*<sub>95%</sub>]<br>*Mdn* (*Min* to<br>*Max*) | -5.4 [-7.4 to -<br>3.3]<br>-5.0 (-15.0 to<br>5.0) | -5.6 [-8.5 to -<br>2.6]<br>-4.5 (-15 to 5) | 0.2 [-2.3 to<br>2.7]<br>0.0 (-7.0 to<br>9.0) | -2.7 [-4.6 to -<br>0.8]<br>-3.0 (-9.0 to<br>4.0) |
| Duration | *M* [*CI*<sub>95%</sub>]<br>*Mdn* (*Min* to<br>*Max*) | -2.9 [-4.1 to -<br>1.6]<br>-2.0 (-8.0 to<br>2.0) | -2.6 [-4.1 to -<br>1.1]<br>-3.0 (-8.0 to<br>4.0) | -0.1 [-1.0 to<br>0.8]<br>0.0 (-4.0 to<br>3.0) | -1.6 [-2.4 to -<br>0.7]<br>-1.0 (-4.0 to<br>1.0) |
| Physical concomitants | *M* [*CI*<sub>95%</sub>]<br>*Mdn* (*Min* to<br>*Max*) | -0.5 [-1.0 to<br>0.1]<br>0.0 (-4.5 to<br>1.5) | -0.8 [-1.8 to<br>0.2]<br>-0.5 (-6.5 to<br>2.5) | 0.3 [-0.7 to<br>1.4]<br>0.0 (-2.0 to<br>7.0) | -0.7 [-1.4 to -<br>0.01]<br>-0.5 (-4.5 to<br>2.0) |
| Subjective parent rating | *M* [*CI*<sub>95%</sub>]<br>*Mdn* (*Min* to<br>*Max*) | -2.75 [-3.9 to<br>-1.6]<br>-3 (-8 to 3) | -3.1 [-4.6 to -<br>1.5]<br>-3 (-7 to 2) | nan | -1.8 [-3.2 to -<br>0.5]<br>-2 (-5 to 2) |

### Stuttering status after Module 2 and after Module 3

According to the SSI-4 severity assessment conducted after Module 2, at T3, 6 children were classified as having no stuttering, 23 as very mild, 5 as mild, and 8 as moderate. Compared to baseline, at T0, stuttering severity decreased by three grades in 1 case, by two grades in 10 cases, and by one grade in 15 cases. In 14 cases, severity remained unchanged, of these, 6 were already classified as very mild. In 2 cases, severity increased by one grade.

Following Module 3, 2 children were classified as having no stuttering, 18 as very mild, 6 as mild, and 2 as moderate. Compared to the end of Module 2 (T3), severity decreased by two grades in 3 cases and by one grade in 4 cases. In 15 cases, severity remained unchanged, of these, 11 were already very mild and 1 had been classified as no stuttering. Severity increased by one grade in 6 cases and by two grades in 1 case.

Finally, 29% of the cases were excluded from Module 3 analysis due to attrition, with 19% of families, that is 8 cases, choosing not to continue treatment. While some level of attrition bias is unavoidable given the structure of the program, it is important to note that most of these children had already demonstrated meaningful progress by the end of Module 2. As shown in Figure 3B (red lines), stuttering severity improved in all but one of the children excluded at this stage. Among the eight families who chose to pause after Module 2, 1 child was classified as having no stuttering, 5 as very mild, 1 as mild, and 1 as moderate. Stuttering severity was reduced in five children by 2 grades, and in 1 child by 1 grad. It remained at a very mild level in 2 children, and was moderate in 1 child (see Supplementary Table 7). Notably, the two children who remained in the very mild range still demonstrated improvement, with SSI-4 total scores decreasing from 9 to 6 and from 4 to 2, respectively. The child with moderate stuttering also showed measurable progress, with the SSI-4 total score decreasing from 25 to 20 and percent stuttered syllables reducing from 12% to 8%. Taken together, these data suggest that the majority of children who discontinued after Module 2, had experienced clinically meaningful improvements in speech fluency.

### Parents’ reports

Parents’ subjective ratings confirmed the positive impact of treatment on their children’s stuttering. After completing Module 2, parents reported a significant reduction in perceived stuttering severity, with an average change score of -2.4 (95% CI [-3.2, -1.6], *T*(31) = -5.82, *p* < .001). This improvement continued following Module 3, with a mean change of -2.6 (95% CI [-3.7, -1.5], *T*(24) = -5.10, *p* < .001).

Among the eight families who discontinued after Module 2, five provided severity ratings via the online survey. Of these, three parents rated their child’s stuttering severity as 1, and two rated it as 2, suggesting continued low perceived severity even without completing the final module.

### Online questionary about coping strategies

Parents’ were also asked to rate their experience across a set of items assessing how they coped with and managed their child’s stuttering. This questionnaire was administered as part of a cohort design and was not controlled, as data were collected at only four time points: baseline (T0), and after each intervention module (T1, T3, T5; see Figure 1A). Figure 3C presents a heatmap illustrating changes in parental ratings from baseline (T0) to the end of Module 2 (T3). Each column represents an individual case, with the rightmost column displaying the average change across all participants.

For the first five items, where higher scores indicate improved coping, positive changes are visualized using shades of red, with darker reds representing greater improvement. This visual approach summarizes the pattern of responses across families. For instance, the Item 1, “*I can handle my child’s stuttering calmly,*” showed an increase in scores for 27 families, no change for 3 families, and a decrease for 3 families. A one sample *t*-test revealed a significant improvement, *T*(32)= 6.18, *p* > 0.001, as indicated by three asterisks in the rightmost cell of this row (also see Supplementary Table 6).

For the remaining items, where lower scores reflect better outcomes, improvements are represented in shades of blue, with darker blues indicating larger reductions in score. For example, Item 8, “*I feel uncomfortable talking to others about stuttering,*” showed a decrease in scores for 12 families, no change for 14 families, and an increase for 6 families. This change was not statistically significant, *T*(32)= -1.38, *p* = 1.0, as denoted by the absence of asterisks.

Overall, most items demonstrated clear improvement across participants. Statistically significant changes were observed in all items except for Item 8, as reflected in the asterisk markers in the average column on the right side of Figure 3C and detailed in Supplementary Table 6.

At the end of Module 1, significant improvements were observed for Items 1 to 3 and Item 5 among the positively worded items, and for Items 6 and 10 among the negatively worded items. These results suggest that early in the intervention, parents began to feel more capable and experienced some relief in their emotional responses related to their child’s stuttering. Following completion of Module 3, the remaining families reported improvements across nearly all items. However, no significant changes were observed for three items: Item 4 (“*I feel confident talking about stuttering with people I trust*.”), Item 8 (“*I feel uncomfortable talking to others about stuttering*”), and Item 9 (“*When my child reacts emotionally to his/her stuttering or talks about it, I often feel overwhelmed*”). These results suggest that while the intervention had a broad impact on parental coping, some aspects, particularly those related to social support and public disclosure, may require additional focus.

Exploratory analyses using Spearman correlations showed no significant associations between changes in stuttering severity (SSI total score) and changes in parental ratings from T0 to T3.

### Interrater-reliability

Interrater agreement was found to be almost-perfect, following the interpretation of Hayes and Krippendorff (Hayes & Krippendorff, 2007). Specifically, Krippendorff’s alpha was 0.88 for the total SSI score and 0.92 for stuttering frequency, indicating strong consistency among raters. These findings were further supported by intraclass correlation coefficients (ICCs), which also reflected high reliability. The ICC for the total SSI score was 0.88, with a 95% CI [0.80, 0.93], while the ICC for stuttering frequency was even higher at 0.92 (95% CI [0.86, 0.96]).

## Discussion

The newly developed parent-child program significantly reduced stuttering, yielding a large effect size within 9 months post-baseline. Our findings provide the first evidence that child-friendly fluency shaping can be feasibly integrated into a parent-child therapeutic framework and effectively delivered to preschoolers. The program’s efficacy appears comparable to outcomes reported in previous randomized controlled trials of stuttering interventions for children aged 3 to 6 years (Harris et al., 2002; Jones et al., 2005; Lattermann et al., 2008; Lewis et al., 2008). The use of a simulated wait-list control helps to mitigate the influence of spontaneous remission, strengthening confidence in treatment effect. Although attrition was relatively high, it is noteworthy that nearly all children whose families paused after Module 2 still demonstrated measurable reductions in stuttering. These key aspects will be discussed in more detail in the sections that follow.

Fluency shaping is a well-established method for treating adults who stutter and seek increased fluency (Euler et al., 2014; Webster, 1980; Wolff-von-Gudenberg & Euler, 2000), and has also shown effectiveness in school-age children (Euler et al., 2021). In preschoolers, however, its use may be more controversial, underscoring the need to monitoring its impact on children’s attitudes toward their speech and overall well-being. Although we did not directly assess the children’s experiences with the speech technique, we gathered parental perceptions at the end of Module 3. Parents were asked to rate, on a scale from 1 to 5, how comfortable they believed their child felt with relaxed phonation and soft speech. Of the families who continued treatment through Module 3, 25 responded via the online survey. One family reported that the child felt very comfortable, 11 reported feeling comfortable, 9 reported partial comfort, 3 reported less comfort, and 1 reported that the child did not feel comfortable at all. These self-reported data, while limited in scope and sample, indicate that some children experienced fluency shaping as comfortable, but that more than half perceived it as only partially or less comfortable. This suggests that while the technique can be acceptable for many, it may not be universally suitable. The findings highlight variability in comfort levels and the importance of tailoring the intervention to individual needs, closely monitoring children’s responses, and further investigating for whom fluency shaping is most beneficial.

The evaluation of a therapeutic approach can be designed in several ways. In our study, we choose the simulated wait-list design, which allowed us to control for the influence of spontaneous remission, a particularly relevant factor in early childhood stuttering (Yairi & Ambrose, 2013). Another common design is to compare a novel or untested intervention with a treatment that has already demonstrated efficacy (Sonneville-Koedoot et al., 2015). Such comparisons can reveal whether the new approach is inferior, equivalent, or superior to the established one. A third design strategy involves comparing two untested interventions, offering the potential to identify which yields better outcomes (Bafrooei et al., 2025). Given that young children exhibit high neuroplasticity and that stuttering follows dynamic developmental courses at this age (Chow et al., 2023; Chow & Chang, 2017), it is especially important to account for spontaneous remission. Therefore, designs that incorporate a control for natural recovery are particularly valuable when evaluating early interventions.

The timing and type of intervention are critical considerations in the treatment of developmental stuttering, particularly with regard to fluency-shaping approaches in preschool-age and school-age children. Our findings suggest that early intervention may yield more favorable outcomes than interventions initiated later in childhood. This advantage may stem from the brain’s heightened neuroplasticity during early childhood, which supports more efficient speech motor learning and adaptation. This neural flexibility, combined with a generally lower emotional burden for both children and their parents at this age, may contribute to the more robust outcomes observed. While these interpretations align with developmental neuroscience (Chang et al., 2025; Chow et al., 2023), they warrant further investigation using neuroimaging methods to examine how early intervention interacts with neural mechanisms of speech production and self-regulation.

In contrast, a recent randomized controlled trial involving German school-age children who stutter found no significant effect on the SSI-4 total score after 3 months (*d* = 0.092) and only a modest improvement after 12 months (Kohmäscher 2023). These results reflect a broader trend in school-age intervention studies, where the focus shifts from fluency enhancement to reducing the negative impact of stuttering on quality of life by addressing their attitude toward stuttering, with moderate improvements *d* = 0.62 (Kohmäscher 2023).

Given these findings, there is a pressing need for well-controlled randomized trials to evaluate the efficacy of fluency-shaping techniques in children. Such studies could help clarify whether the limited effects observed to date are due to reduced neuroplasticity, suboptimal timing, or mismatches between the intervention approach and developmental stage. Our own findings, while preliminary, emphasize the potential benefits of early fluency-based treatment and underscore the importance of optimizing both timing and method. Similar benefits observed in early indirect approaches, such as those reported in the RESTART-DCM (Sonneville-Koedoot et al., 2015) further support the case for early, well-targeted interventions. However, the complex relationship between intervention timing, treatment type, and long-term psychosocial outcomes remains an open question and requires further study (Leclercq et al., 2024).

A key challenge in evaluating interventions for preschoolers who stutter is the high probability of spontaneous remission (Reilly et al., 2009; Yairi & Ambrose, 2013). As noted earlier, previous prospective controlled trials with both school-age (Craig et al., 1996; Euler et al., 2021; Kohmäscher et al., 2023) and preschool-age children (Harris et al., 2002; Jones et al., 2005; Lattermann et al., 2008; Lewis et al., 2008) have addressed this confound using wait-list control designs to help distinguishing treatment effects from spontaneous recovery. Following this approach, our study simulated a parallel wait-list control group that did not receive direct intervention until approximately six months after baseline. At study entry, both groups had comparable stuttering durations since stuttering onset (*Mdn* = 20 months). While the early intervention group showed a significant reduction by 9 months post-baseline, after completing Module 2, the wait-list group showed no comparable improvement during this same period (See Figure 3A). This strengthens the interpretation that the observed improvements were primarily attributable to the intervention. However, we acknowledge that individual recovery potential may interact with treatment outcomes, and we do not suggest that intervention alone accounts for all observed improvements.

Our data offer only a limited snapshot of the developmental trajectory of assisted stuttering remission, limited to a three-month intervention window. During this period, children who continued beyond Module 1 showed notable improvements in fluency. Still, the fluctuating nature of early stuttering is evident, with some children showing both improvement and later regression in SSI-4 scores across T0, T3, and T5. Although the intervention extended over 12 months, this does not constitute long-term follow-up, and we are therefore cautious in making definitive classifications of remission or persistence at T5. In some cases, stuttering severity increased between T3 and T5 despite earlier gains. To avoid overinterpreting these short-term fluctuations, we have chosen not to stratify the data by remission status, as such distinctions would require longer-term follow-up data to be reliable.

Having said this, we consider the findings of the current study preliminary as the retrospective design did not allow for random assignment to early treatment or wait-list-control groups, introducing potential selection bias. We controlled for variables, such as sex, age, stuttering onset, time since onset, and baseline severity to minimize such biases. Observer bias is also a concern, as SSI-4 raters knew that all participants received treatment, possibly leading to favorable interpretations of ambiguous cases. To address this, we simulated a waitlist group and blinded raters for the timing of the speech samples. The observed improvement in the early group after 9 months, with no change in the delayed group, supports the efficacy of the treatment. Additionally, since participants and parents knew they were receiving treatment, placebo effects or response bias could have affected subjective ratings, although the objective SSI-4 measure is less prone to these biases.

A key strength of this study lies in its use of a retrospective simulated wait-list design, which, to our knowledge, has not previously been applied in early childhood stuttering research. This approach offers the ethical and practical advantages of a wait-list comparison without requiring families to knowingly delay access to treatment. By leveraging pre-existing clinical data and collecting outcome measures prior to group assignment, the design avoids potential demotivation, dropout, and dissatisfaction often associated with prospective wait-list controls. Importantly, the study was conducted within a real-world clinical setting, reflecting how the intervention is delivered in everyday practice. This enhances the ecological validity of the findings and underscores the feasibility of applying the method in typical service contexts. In clinical scenarios where timely intervention is critical, such as early childhood stuttering, this methodology offers a promising framework for evaluating treatment efficacy while minimizing ethical concerns. We believe it holds potential for broader use in developmental and pediatric intervention research.

### Limitations

A key limitation of this study is the absence of direct outcome measures assessing the emotional and psychological effects of the intervention in children. We cannot draw conclusions about the long-term emotional impact of the therapy, particularly for those children who continued to stutter after treatment. Although the primary focus was on speech fluency and parent-reported coping, this leaves important questions unanswered regarding how children themselves experience and internalize fluency-shaping interventions.

Given concerns that such approaches may inadvertently influence children’s attitudes or self-perception, especially in those for whom stuttering persists, future research should incorporate validated measures of emotional, social, and cognitive outcomes such as the Communication Attitude Test for Preschoolers and Kindergartners (KiddyCat, (Vanryckeghem et al., 2005) and the Overall Assessment of the Speaker’s Experience of Stuttering for School-age children (OASES-S, (Yaruss et al., 2012; Yaruss & Quesal, 2014) to more fully assess the broader impact of intervention.

Furthermore, the study design did not allow for systematic long-term follow-up of stuttering outcomes at later time points such as T5. As such, we are unable to report on the persistence or relapse of stuttering beyond the initial post-treatment phase. This is an important limitation in assessing the sustained efficacy of Frankini. Future studies, particularly prospective randomized controlled trials, should include longitudinal follow-ups and measures of maintenance or change in stuttering severity over time. These should be combined with child-centered assessments to ensure that both objective and subjective outcomes are adequately captured, ultimately providing a more comprehensive evaluation of treatment effectiveness.

Another limitation of the present study is the potential for selection bias related to differences between children who began therapy earlier versus those in the delayed treatment group. As the study was retrospective, we could not systematically capture the reasons behind delayed therapy initiation. Importantly, the intervention was delivered in a group format with limited capacity, and logistical constraints likely influenced enrollment timing. For example, some families may have experienced delays because courses were already fully booked, while in other cases, the institution had to postpone starting a group until a minimum number of participants had registered. These factors suggest that delayed therapy was not necessarily a reflection of lower parental motivation or child-related variables. Nevertheless, we cannot rule out the possibility that differences in family engagement, help-seeking behavior, or other unmeasured factors contributed to treatment timing and outcomes. Future prospective studies should aim to control for such potential confounders or use randomization to ensure more balanced group comparisons.

While some degree of attrition bias was unavoidable due to the program’s flexible design, stuttering severity improved for all but one of the children who discontinued after Module 2. These improvements are consistent with parental ratings available for five of these children, suggesting that attrition likely did not systematically bias the results toward more favorable outcomes. As this study is based on retrospectively recorded clinical data, outcome measures are inherently vulnerable to missing data due to participant dropout. In future prospective studies, this limitation could be addressed by informing families from the outset about the importance of completing follow-up assessments at multiple time points, even if treatment is paused or ended early. This approach would help ensure more complete and systematic outcome data.

### Clinical implications and recommendations for practitioners

The findings of this study have several important implications for clinical practice. First, the observed benefits of early intervention support current guidelines that emphasize timely referral and treatment initiation in preschool-aged children who stutter (Brignell et al., 2021; Keilmann et al., 2018; K. Neumann et al., 2016; Sjøstrand et al., 2021). The results also suggest that even brief delays in therapy access may impact outcomes, reinforcing the need for efficient resource allocation and reduced waiting times.

In line with previous research, it is important to consider how the present approach compares with other established therapy methods for this age group, such as the Lidcombe Program, Palin Parent-Child Interaction therapy, and the RESTART Demands and Capacities Model. Evidence from controlled trials indicates that these approaches can also yield positive outcomes in preschool children who stutter (e.g., (Millard et al., 2009; Onslow et al., 1994; Sonneville-Koedoot et al., 2015). While direct comparisons between the current intervention and these methods are not yet available, the present findings suggest that the Frankini method may be a viable alternative, particularly in settings where structured group formats and strong parental involvement are feasible. Importantly, not all programs are equally available across countries (for example, some approaches are more accessible in Germany, while others are not), and clinical choices must therefore be adapted to local service structures and training opportunities.

Accordingly, clinical decision-making should take into account not only treatment efficacy but also contextual factors, such as therapist training, family preferences, and service delivery structures. At this stage, the available evidence does not allow us to conclude that one method is uniformly superior. Rather, our results highlight the importance of offering timely, accessible, and family-centered interventions, of which the Frankini method represents one promising option.

Finally, the use of parent-reported coping outcomes highlights the importance of involving families in the therapeutic process (Bergþórsdóttir et al., 2022) and considering both objective and subjective indicators of progress (Onslow et al., 2018). Clinicians may also consider how structured group formats can be effectively implemented within clinical settings while balancing logistical constraints.

### Conclusion

Early intervention is crucial as it targets stuttering before it becomes deeply ingrained, leading to improved long-term outcomes and reducing its impact on the child’s overall quality of life. Our novel parent-child program effectively reduced stuttering severity in preschoolers, while enhancing parents’ attitudes and coping strategies. Future research should include follow-up assessments to determine the long-term sustainability of the intervention’s benefits along with measures of children’s self-perception of their speech and communication abilities.

## Data Availability

All data produced in the present work are contained in the manuscript

## Acknowledgments

This project was supported by the University Medical Center Göttingen, Germany. IN, AM, KA, and KA have received institutional support from KST GmbH for the development of the parent-child program.

## Data Availability Statement

Owing to ethical concerns, supporting data cannot be made openly available. Further information regarding the data and conditions for access is available from the corresponding author.

## Notes

### Competing Interest Statement

Imke Niemann, Anna Merkel, Kristina Anders, Katja Hente, Alexander Wolff von Gudenberg reports financial support was provided by KST Institut GmbH.

### Funding Statement

This project was supported by the University Medical Center Goettingen, Germany.

### Author Declarations

Ethics committee/IRB of the University Medical Center Goettingen gave ethical approval for this work. Ethic-Nr. 4/5/24

### Summary of Updates

We have revised the manuscript. In particular, we have revised the Abstract, Highlights, Introduction, Methods, Results, Discussion, and Clinical Implications sections to: (1) clarify the primary aim of RESTART-DCM and update the corresponding reference, (2) amend the conceptual framing of the Frankini method to clarify that it aligns with a neurodiversity perspective by fostering acceptance and resilience, (3) elaborate further on the comparison of the Frankini method with other treatment methods for preschool-aged children, (4) and to emphasize the benefits children experience from early treatment and highlight the need for future studies to monitor the psycho-emotional impact of the method in greater depth.

