## Supplementary for "Efficacy of a parent-child program for 3- to 6-year-old children with stuttering: a retrospective controlled wait-list group pilot study"

**Supplemental information**

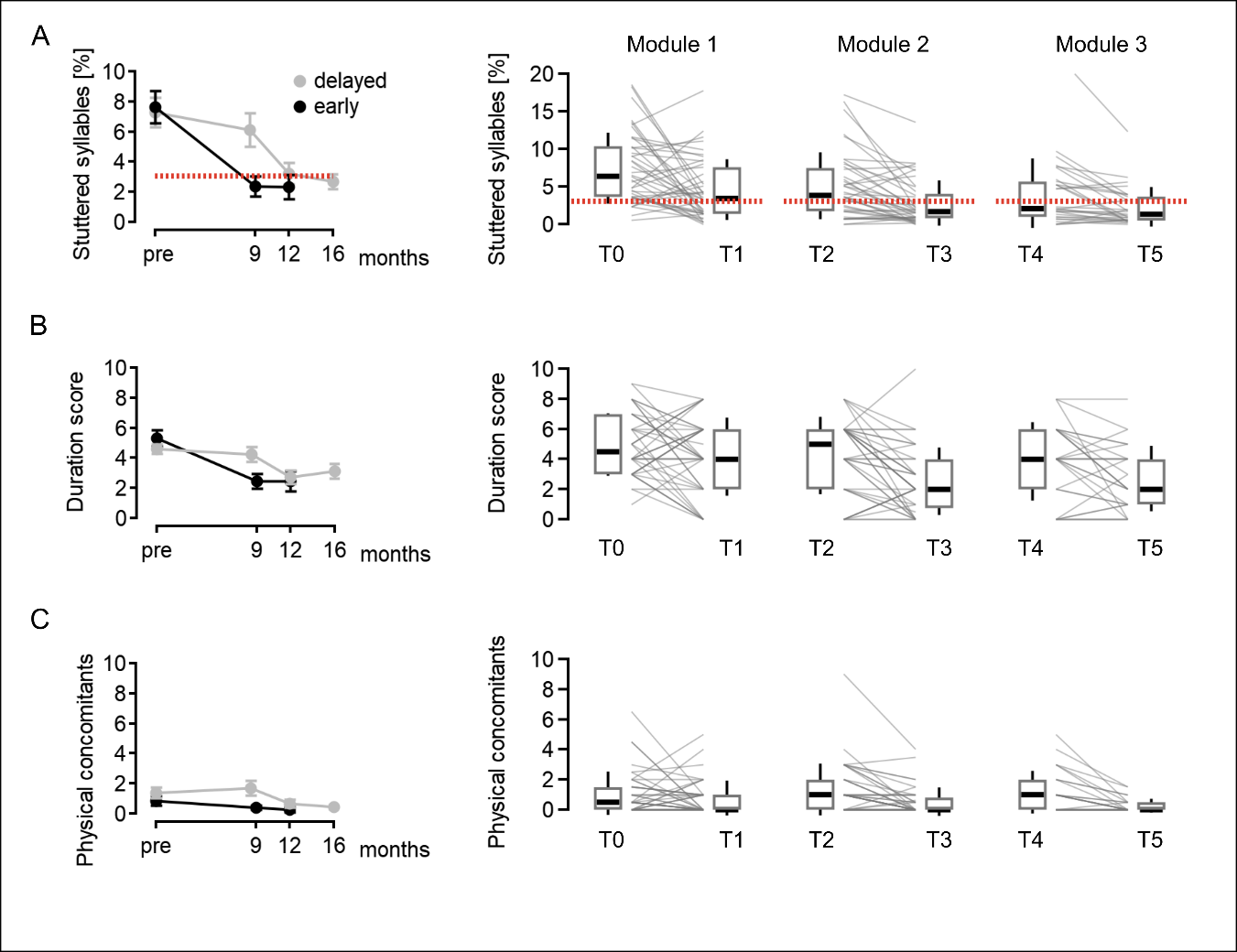

**Supplementary Figure 1**. **Stuttering severity.** A) Percent stuttered syllables, B) average duration of the three longest stuttering events, and C) physical concomitants. The first plot in a row depicts group averages and standard errors separated for the early and delayed group. The following box plots include individual curves separated per module. The dotted red lines indicate the cut-off vale for stuttering.

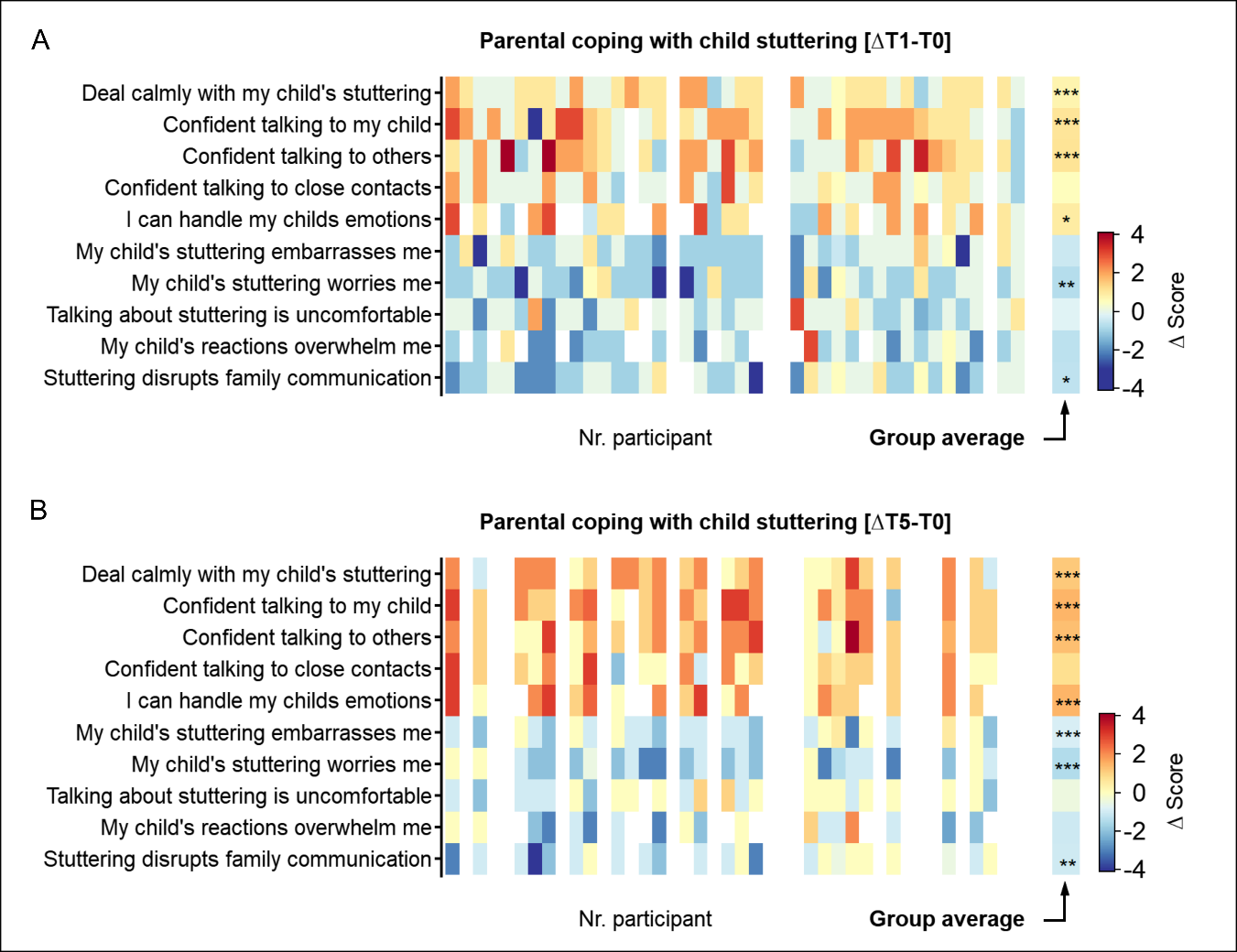

**Supplementary Figure 2.** **Changes in parental experience with their child's stuttering** A) between baseline and end of Module 1, and B) between baseline and end of Module 3. White cells indicate missing data. The last column indicates the grand mean for a given item with **p* < .05, ***p* < .01, and ****p* < .001 (two-sided one sample *t*-test).

**Supplementary Table 1. Baseline characteristics**

| **Characteristic** | **Early-treatment**  *n* = 23 | **Delayed-treatment**  *n* = 19 | **Difference**  *T*(*df*)  *p* |
| --- | --- | --- | --- |
| Age diagnostics (T0), mo  *M* (*SD*)  *Mdn* (*IQR*)  *Min*-*Max* | 52.7 (8.8)  54.9 (10.6)  35.1-69.5 | 51.6 (7.6)  54.1 (11.9)  36.0-63.1 | 0.44 (40.0)  .663 |
| Age start presence 1 (T2), mo  *M* (*SD*)  *Mdn* (*IQR*)  *Min*-*Max* | 58.0 (8.7)  59.8 (11.0)  40.2 - 74.4 | 59.7 (7.7)  59.7 (12.5)  44.7 - 71.9 | -0.66 (39.8)  .511 |
| Δ T3 - T0, mo  *M* (*SD*)  *Mdn* (*IQR*)  *Min*-*Max* | 9.0 (0.6)  9.0 (0.7)  6.9 - 9.8 | 11.7 (1.0)  11.7 (1.4)  10.0 - 12.9 | -10.8 (30.1)  **< .001** |
| Δ T5 - T0, mo  *M* (*SD*)  *Mdn* (*IQR*)  *Min*-*Max* | 12.8 (1.9)  12.4 (0.7)  10.9 - 19.6 | 15.9 (2.1)  15.6 (2.2)  13.2 - 20.4 | -4.56 (28.8)  **< .001** |
| Family history  None (*n*)  First-degree-relative (*n*)  Second-degree-relative (*n*)  Third-degree-relative (*n*)  Fourth-degree-relative (*n*)  Unknown (*n*) | 7 (30.4%)  6 (26.1%)  4 (17.4%)  2 (8.7%)  2 (8.7%)  2 (8.7%) | 7 (36.8%)  6 (31.6%)  4 (21.1%)  0  2 (10.5%)  0 |  |
| Monolingual German (*n*)  Bilingual (*n*) | 16 (69.6%)  7 (30.4%) | 16 (84.2%)  3 (15.8%) |  |
| Former treatment  None (*n*)  Logopedics (*n*)  MiniKids (*n*)  Lidcombe (*n*)  Van Riper (*n*) | 12 (52.2%)  9 (39.1%)  1 (4.3%)  1 (4.3%)  0 | 8 (42.1%)  6 (31.6%)  2 (10.5%)  2 (10.5%)  1 (5.3%) |  |

**Supplementary Table 2. Linear mixed model revealed a group by time interaction (SSI total score)**

| SSI total | **Contrast** | **Linear mixed-effects model fit by maximum likelihood**  SSI ~ 1 + age + sex, random = ~ 1\| subject / time  + group  + time  + group × time | | | | | | **ANOVA** | |
| --- | --- | --- | --- | --- | --- | --- | --- | --- | --- |
|  |  | *B* | *SE* | 95% fw. *CI* | *Z* | Adj. *p** | r | *χ²*(*df*) | *p* |
| (intercept) |  | 16.87 | 5.58 | [2.48, 31.25] | 3.13 | **.009** |  |  |  |
| Age |  | -0.02 | 0.09 | [-0.26, 0.22] | -0.18 | 1 |  |  |  |
| Sex |  | -0.09 | 0.80 | [-2.16, 1.98] | -0.12 | 1 |  |  |  |
| Group | Early < delayed | -0.32 | 1.09 | [-3.12, 2.48] | -0.31 | 1 |  | 5.07 (7) | **.024** |
| Time | 9 mo < baseline | 4.24 | 1.38 | [0.67, 7.81] | 3.17 | **.009** |  | 21.95 (9) | **<.001** |
|  | 12 mo < baseline | 7.26 | 1.46 | [3.49, 11.03] | 5.15 | **< .001** |  |  |  |
| Group × Time | Early < delayed × 9 mo < basel. | -4.50 | 1.38 | [-8.07, -0.93] | -3.37 | **.005** | .36 | 10.61 (11) | **.005** |
|  | Early < delayed × 12 mo < basel. | -2.16 | 1.46 | [-5.93, 1.61] | -1.53 | 1 | .17 |  |  |

**adjusted p values; holm method*

**Supplementary Table 3. Linear mixed model revealed a group by time interaction (disfluent syllables %)**

| Stuttered  syllables % | **Contrast** | **Linear mixed-effects model fit by maximum likelihood**  StutFreq ~ 1 + age + sex, random = ~ 1\| subject / time  + group  + time  + group × time | | | | | | **ANOVA** | |
| --- | --- | --- | --- | --- | --- | --- | --- | --- | --- |
|  |  | *B* | *SE* | 95% fw. *CI* | *Z* | Adj. *p** | r | *χ²*(*df*) | *p* |
| (intercept) |  | 5.91 | 3.79 | [-3.89, 15.71] | 1.62 | .531 |  |  |  |
| Age |  | 0.02 | 0.06 | [-0.14, 0.19] | 0.36 | 1 |  |  |  |
| Sex |  | -0.28 | 0.55 | [-1.69, 1.14] | -0.53 | 1 |  |  |  |
| Group | Early < delayed | -0.35 | 0.65 | [-2.03, 1.32] | -0.56 | 1 |  | 1.31 (7) | .252 |
| Time | 9 mo < baseline | 3.05 | 0.70 | [1.23, 4.87] | 4.49 | **>.001** |  | 33.50 (9) | **<.001** |
|  | 12 mo < baseline | 4.68 | 0.75 | [2.74, 6.61] | 6.47 | **>.001** |  |  |  |
| Group × Time | Early < delayed × 9 mo < basel. | -2.21 | 0.70 | [-4.03, -0.39] | -3.26 | **.007** | .35 | 10.01 (11) | **.007** |
|  | Early < delayed × 12 mo < basel. | -0.92 | 0.75 | [-2.86, 1.01] | -1.28 | .804 | .14 |  |  |

**adjusted p values; holm method*

**Supplementary Table 4. Linear mixed model revealed a group by time interaction (SSI frequency)**

| **Frequency** | **Contrast** | **Linear mixed-effects model fit by maximum likelihood**  frequency ~ 1 + age + sex, random = ~ 1\| subject / time  + group  + time  + group × time | | | | | | **ANOVA** | |
| --- | --- | --- | --- | --- | --- | --- | --- | --- | --- |
|  |  | *B* | *SE* | 95% fw. *CI* | *Z* | Adj. *p** | r | *χ²*(*df*) | *p* |
| (intercept) |  | 10.34 | 3.82 | [0.48, 20.21] | 2.81 | **.025** |  |  |  |
| Age |  | -0.009 | 0.06 | [-0.17, 0.16] | -0.14 | 1 |  |  |  |
| Sex |  | -0.20 | 0.55 | [-1.62, 1.23] | -0.37 | 1 |  |  |  |
| Group | Early < delayed | -0.21 | 0.66 | [-1.91, 1.48] | -0.34 | 1 |  | 4.37 (7) | **.037** |
| Time | 9 mo < baseline | 2.59 | 0.72 | [0.74, 4.44] | 3.74 | **.001** |  | 24.77 (9) | **<.001** |
|  | 12 mo < baseline | 4.05 | 0.76 | [2.08, 6.03] | 5.51 | **<.001** |  |  |  |
| Group × Time | Early < delayed × 9 mo < basel. | -2.80 | 0.72 | [-4.66, -0.95] | -4.04 | **<.001** | .42 | 14.83 (11) | **<.001** |
|  | Early < delayed × 12 mo < basel. | -1.37 | 0.76 | [-3.34, 0.60] | -1.86 | .251 | .21 |  |  |

**adjusted p values; holm method*

**Supplementary Table 5. Linear mixed model revealed a group by time interaction (SSI duration)**

| **Duration** | **Contrast** | **Linear mixed-effects model fit by maximum likelihood**  duration ~ 1 + age + sex, random = ~ 1\| subject / time  + group  + time  + group × time | | | | | | **ANOVA** | |
| --- | --- | --- | --- | --- | --- | --- | --- | --- | --- |
|  |  | *B* | *SE* | 95% fw. *CI* | *Z* | Adj. *p** | r | *χ²*(*df*) | *p* |
| (intercept) |  | 5.97 | 2.17 | [0.34, 11.59] | 2.84 | **.022** |  |  |  |
| Age |  | -0.02 | 0.04 | [-0.11, 0.07] | -0.56 | 1 |  |  |  |
| Sex |  | 0.01 | 0.31 | [-0.80, 0.83] | 0.05 | 1 |  |  |  |
| Group | Early < delayed | -0.48 | 0.36 | [-1.40, 0.44] | -1.39 | .491 |  | 0.25 (7) | .615 |
| Time | 9 mo < baseline | 1.46 | 0.35 | [0.55, 2.37] | 4.29 | **<.001** |  | 29.45 (9) | **<.001** |
|  | 12 mo < baseline | 2.21 | 0.38 | [1.23, 3.18] | 6.08 | **<.001** |  |  |  |
| Group × Time | Early < delayed × 9 mo < basel. | -1.41 | 0.35 | [-2.32, -0.50] | -4.13 | **<.001** | .42 | 15.49 (11) | **<.001** |
|  | Early < delayed × 12 mo < basel. | -0.63 | 0.38 | [-1.60, 0.34] | -1.73 | .334 | .19 |  |  |

**adjusted p values; holm method*

**Supplementary Table 6. One-sample *t*-tests indicate change in parental coping with child stuttering**

|  | **ΔT1-T0** | | **ΔT3-T0** | | **ΔT5-T0** | |
| --- | --- | --- | --- | --- | --- | --- |
|  | *T* (*df*) | *p^a^* | *T* (*df*) | *p^a^* | *T* (*df*) | *p^a^* |
| I can handle my child's stuttering calmly. | 5.62 (37) | <.001 | 6.18 (32) | <.001 | 5.50 (24) | <.001 |
| I feel confident talking to my child about stuttering. | 5.37 (36) | <.001 | 9.59 (31) | <.001 | 6.31 (23) | <.001 |
| I feel confident talking about stammering with less familiar people. | 4.90 (36) | <.001 | 5.69 (30) | <.001 | 5.50 (23) | <.001 |
| I feel confident talking about stuttering with people I trust. | 3.18 (36) | .091 | 4.12 (30) | .008 | 3.20 (23) | .119 |
| I can handle my child's emotions and comments about his/her stuttering well. | 3.75 (26) | .027 | 5.11 (22) | .001 | 6.07 (18) | <.001 |
| My child's stuttering embarrasses me. | -2.97 (37) | .156 | -4.70 (31) | .002 | -5.12 (24) | <.001 |
| I am concerned about my child's stuttering. | -4.50 (37) | .002 | -7.24 (32) | .001 | -7.60 (24) | <.001 |
| I feel uncomfortable talking to others about stuttering. | -1.07 (36) | 1 | -1.38 (31) | 1 | -2.70 (23) | .387 |
| When my child reacts emotionally to his/her stuttering or talks about it, I often feel overwhelmed. | -3.16 (27) | .116 | -3.80 (21) | .031 | -3.23 (17) | .143 |
| Occurring stuttering disrupts communication in our family. | -3.75 (36) | .019 | -4.05 (30) | .01 | -4.56 (22) | .005 |

*^a^Bonferroni correction for multiple comparison involving 30 tests*

**Supplementary Table 7. Demographic characteristics and outcome measures for participants who discontinued the program after completing Module 2.**

| **ID** | **Group** | **Sex** | **Age onset** | **Age baseline** | **ΔAge baseline - onset** | **Age start M1** | **SSI  T0** | **SSI T3** | **ΔSSI T3 - T0** | **Stutt. freq. T0** | **Stutt. freq. T3** | **ΔStutt. freq. T3 - T0** | **SSI-4 severity T0** | **SSI-4 severity T3** |
| --- | --- | --- | --- | --- | --- | --- | --- | --- | --- | --- | --- | --- | --- | --- |
| sub50 | late | male | 36 | 61 | 25 | 64 | 12 | 0 | -12 | 2.4 | 0.6 | -1.8 | mild | no stuttering |
| sub13 | early | male | 38 | 47 | 9 | 47 | 22 | 5 | -17 | 9.1 | 1.0 | -8.2 | moderate | very mild |
| sub16 | early | male | 30 | 39 | 9 | 40 | 21 | 4 | -17 | 6.3 | 0.8 | -5.6 | moderate | very mild |
| sub64 | late | male | 29 | 57 | 28 | 61 | 17 | 8 | -10 | 0.3 | -4.2 | -4.2 | moderate | very mild |
| sub63 | late | male | 24 | 54 | 30 | 58 | 20 | 11 | -9 | 13.8 | 3.7 | -10.2 | moderate | mild |
| sub15 | early | male | 29 | 39 | 10 | 39 | 9 | 6 | -4 | 2.9 | 0.9 | -2.0 | very mild | very mild |
| sub07 | late | female | 24 | 56 | 32 | 58 | 4 | 2 | -2 | 0.5 | 0.5 | 0.0 | very mild | very mild |
| sub61 | early | male | 36 | 55 | 19 | 56 | 25 | 20 | -5 | 11.7 | 7.7 | -4.0 | moderate | moderate |

*Age is reported in months*

**Supplementary Table 8. Group assignment, bilingualism, family history, and former treatments**

| **ID** | **Sex** | **Group** | **L1_1** | **L1_2** | **Family history** | **Former therapy** |
| --- | --- | --- | --- | --- | --- | --- |
| sub12 | m | early | Russian | German | cousin | logopedics |
| sub13 | m | early | German |  | none | none |
| sub14 | m | early | German | Polish | father | Lidcombe |
| sub15 | m | early | German | Persian | father, grandfather | none |
| sub16 | m | early | German |  | cousine, uncle | none |
| sub17 | w | early | German |  | great uncle | none |
| sub19 | m | early | German |  | great cousin | none |
| sub22 | m | early | German |  | cousin | logopedics |
| sub23 | w | early | German |  | father, uncle | none |
| sub27 | w | early | German |  | father, uncle, great uncle | logopedics |
| sub32 | m | early | German | Russian | father | logopedics |
| sub33 | m | early | German |  | father, grandmother | logopedics |
| sub43 | m | early | German |  | unknown | MiniKids |
| sub44 | w | early | German | Polish | uncle,nephew, grandmother | none |
| sub45 | w | early | German |  | none | logopedics |
| sub46 | m | early | German |  | unknown | none |
| sub54 | m | early | German |  | uncle | none |
| sub57 | w | early | German | English | none | logopedics |
| sub59 | w | early | English | German | uncle | logopedics |
| sub61 | m | early | German |  | none | logopedics |
| sub65 | m | early | German |  | none | none |
| sub73 | m | early | German |  | none | none |
| sub74 | m | early | German |  | none | none |
| sub02 | w | delayed | German |  | father, uncle, grandfather | none |
| sub03 | m | delayed | German |  | father, grandfather | Lidcombe |
| sub07 | w | delayed | German |  | sister with remission | none |
| sub09 | w | delayed | German |  | great- and great grandfather | logopedics |
| sub10 | m | delayed | German |  | none | MiniKids |
| sub11 | m | delayed | German |  | father, cousin | none |
| sub18 | m | delayed | Albanian | German | great uncle, great aunt | none |
| sub20 | m | delayed | German |  | none | none |
| sub25 | m | delayed | German |  | none | logopedics |
| sub31 | m | delayed | German |  | none | logopedics |
| sub34 | w | delayed | German |  | great cousin | none |
| sub36 | m | delayed | German |  | grandmother | Lidcombe |
| sub37 | m | delayed | German |  | none | MiniKids |
| sub48 | w | delayed | German | Polish | none | logopedics |
| sub50 | m | delayed | German |  | father | logopedics |
| sub51 | w | delayed | German |  | grandmother | none |
| sub55 | m | delayed | German | Persian | aunt | logopedics |
| sub63 | m | delayed | German |  | father | Van Riper |
| sub64 | m | delayed | German |  | none | none |
